# A Non-Pharmacological, Nociceutical Formulation Lessens Chemotherapy-Induced Peripheral Neuropathy in Cancer Patients

**DOI:** 10.1101/2024.12.29.24319628

**Authors:** Sonia Servitja, Maria Castro-Henriques, Iñaki Álvarez-Busto, Carlota Díez-Franco, Alba Medina-Castillo, Maria Asunción Algarra-García, Elena López-Miranda, Margaret Lario-Martínez, Maria Isabel Luengo-Alcázar, Miguel Borregón, Ana Davó, Anna Gassull-Delgado, Sara Roque-García, Ana Gonzaga-López, Jesus Manuel Poveda-Ferriols, Severine Pascal, Clotilde Ferrándiz-Huertas, Ana María Mitroi-Marinescu, Marta García-Escolano, Asia Fernández-Carvajal, Antonio Ferrer-Montiel

## Abstract

**Purpose:** Up to 80% of patients undergoing taxanes or platinum-based chemotherapy (CT) develop a disturbing peripheral polyneuropathy referred to as CIPN, that affects their treatment compliance to CT and long-term quality of life (QoL). Cumulative evidence shows that taxanes and platinum agents sensitize epidermal nociceptive terminals by potentiating the activity of nociceptor thermosensitive channels. Our aim was to evaluate the efficacy and safety of a non-pharmacological nociceutical formulation acting on epidermal nociceptive endings preventing, delaying and/or lessening CIPN sensory symptoms during CT.

**Methods:** We designed a proof-of-concept, double-blind, randomized, two-arms multicenter clinical study (NCT06733545). Participants started a daily topical application of the assigned formulation in hands (moisturizing or nociceutical). Upon appearance of neuropathic symptoms in hands and/or feet, they applied the creams twice daily in hands and feet. Diagnosis and follow up of CIPN grade and adverse effects were conducted by study investigators, as well as a QoL questionnaire.

**Results:** A cohort of 142 patients treated with taxanes and/or platinum agents were randomly assigned to the two groups. Withdrawals were similar in both arms (9 and 14), leading to a balanced number of patients per group (61 moisturizing vs 58 nociceutical). Overall, a similar number of participants developed a peripheral neuropathy in both arms (73% moisturizing vs 67% nociceutical, p=0.1). A lower CIPN incidence in hands was observed in the nociceutical arm (32% vs 13%, p=0.03). Furthermore, the nociceutical formulation delayed the appearance of neuropathic symptoms as compared to the moisturizing cream (6 vs 8 cycle, p=0.009). The Leonard scale questionnaire revealed that the nociceutical formulation attenuated the severity of patients’ neuropathic symptoms from extremely to hardly any (58% vs. 35%, p<0.0017), increasing patient QoL.

**Conclusion:** This pilot study suggests that topical protection of nociceptive epidermal terminals with a topical nociceutical formulation reduced the incidence of CIPN in hands, delayed its onset and increased the QoL of patients. These findings provide solid evidence for a larger, confirmatory clinical study.

## INTRODUCTION

Chemotherapy-induced peripheral neuropathy (CIPN) is a very common side effect that may affect up to 80% of cancer patients treated with chemotherapeutic drugs such as taxanes, platinum agents^1^. CIPN is characterized by disturbing sensory symptoms, including pain, dysesthesia, paresthesia and motor deficits that notably affect the quality of life (QoL) of patients and may lead to changes in the chemotherapeutic treatment, from dose reduction to treatment cessation ^1^. CIPN commonly appears around the 4-6 chemotherapy cycle and aggravate with cumulative doses of chemotherapeutic drugs^2^. Furthermore, CIPN may chronify and last for ≥6 months after chemotherapy in up to 30-40% of patients^3^. Despite its impact in the patient QoL and CT treatment, there are not therapeutic strategies that prevent or ameliorate this condition. Low grade (I/II) CIPN is left without treatment or occasionally managed with sensitive skin creams, while high grade (III/IV) is palliatively treated with duloxetine, gabapentin or Qutenza patches containing 8% capsaicin to reduce the neuropathic symptoms^4^. As preventive measures, for some CT regimes cold gloves are used to limit the amount of CT reaching the distal epidermal terminals by promoting vasoconstriction^4^, along with physical exercise and physiotherapy. Although the scientific support for these strategies is variable.

A plethora of clinical studies have been performed to test different interventions aimed to reduce the incidence of the neuropathy and to mitigate the disturbing sensory symptoms ^5^. These studies encompass: (i) evaluating exercise and resistance training^4^; (ii) cryotherapy and compression theraphy^6^; (iii) wireless cutaneous nerve stimulation^7^; (iv) acupuncture^8^; (v) intravenous monosialotetrahexosylganglioside (GM-1)^9^; (vi) omega-3 fatty acids and essential oils^10,11^; (vii) oral lafutidine^12^; and others (https://clinicaltrials.gov/). Among these studies, exercise has proven beneficial improving patient quality of life^4^. Most of the other have shown some potential benefits for supporting care although they need larger confirmatory studies for solid conclusions^13^. In this regard, the ASCO guidelines summarize all the interventions tested thus far and recommends those that have sufficient clinical support to be offered to patients suffering CIPN, mainly, duloxetine, gabapentin and Qutenza patches^13^. Thus, clinical management of CIPN is yet un unmet need to increase supportive care and patients’ QoL, and to improve treatment compliance^14^.

Pre-clinical evidence shows that CIPN results from the sensitization of distal epidermal sensory terminals resulting from a direct action of chemotherapeutics on sensory endings and on cutaneous and immune cells^15^. Furthermore, cumulative interaction with Schwann cells leads to myelinization deficits and chronification of the neuropathy ^16^. These cellular alterations notably potentiate nociceptor excitability, promoting spontaneous electrical activity^17^. Several studies have signaled to thermosensory channels TRPV1, TRPM8 and TRPA1, along with voltage gated Nav channels, as molecular targets to protect epidermal nociceptive terminals from algesic/pro-inflammatory sensitization^17–19^. Indeed, exposure of sensory neurons *in vitro* to paclitaxel or oxaliplatin for 24h notably increases their spontaneous and evoked electrical activity^17^. This increment in nociceptor excitability is significantly mediated by an increase in the expression and activity of the TRPV1 polymodal sensory channel, along with and alteration of Nav currents^17^. Taken together, these findings suggests that topical protection of nociceptive epidermal terminals may provide a valuable strategy to prevent or delay the onset of CIPN and reduce its severity ameliorating the impact on patient QoL.

Nociceuticals are non-pharmacological moisturizing formulations incorporating active ingredients that protect nociceptive epidermal endings, promoting their physiological activity through a multitarget action that helps to reduce the excitability underlying the sensory neuropathic symptoms. We developed a family of soft molecules that reacted with thermosensory epidermal channels and receptors^20,21^. These molecules were designed with an esterase-sensitive ester group that is hydrolyzed in the dermis preventing a systemic distribution and their accumulation in the skin^20^. Application of the most potent compound reduced thermal hyperalgesia and itch^21^. Thus, these findings suggest that this family of soft compounds could be useful to topically manage CIPN sensory symptoms.

To test this hypothesis, we designed a proof-of-concept, double-blind, randomized, two-arms multicenter clinical study to investigate the protective activity of the nociceutical ingredient Calmapsin^®^, a vanilloid-based, non-pungent soft compound^20^. For this purpose, we evaluated the safety and efficacy of a non-pharmacological nociceutical formulation, that combined Calmapsin^®^ with lipids and tocopherol, preventing, delaying and/or lessening palmoplantar neuropathic sensory symptoms during CT. As comparison, we used the moisturizing base formulation. Our findings suggest that the use of the nociceutical formulation during CT increased the QoL of patients by delaying the onset of CIPN and alleviating the sensory neuropathic symptoms suggesting that protection of epidermic sensory endings may provide a strategy for supportive care and QoL of patients. This observation warrants a larger, confirmatory clinical study.

## PATIENTS AND METHODS

### Patients and study design

This proof-of-concept clinical study was a multicenter, double-blind, randomized, placebo-controlled research project performed in 9 hospitals (Clinical Trials registration NCT06733545) The Ethics Committees of all participating Hospitals approved the protocol and the informed consent documents. At each hospital, medical oncologists acting as principal investigators were responsible for: (i) recruiting volunteer; (ii) checking that they met the inclusion and exclusion criteria (Table 1); (iii) obtaining the signed informed consent; (iv) completing the data collection booklet; (v) and collecting the appearance of CIPN and CIPN grade, along with patient follow up.

**Table 1.**
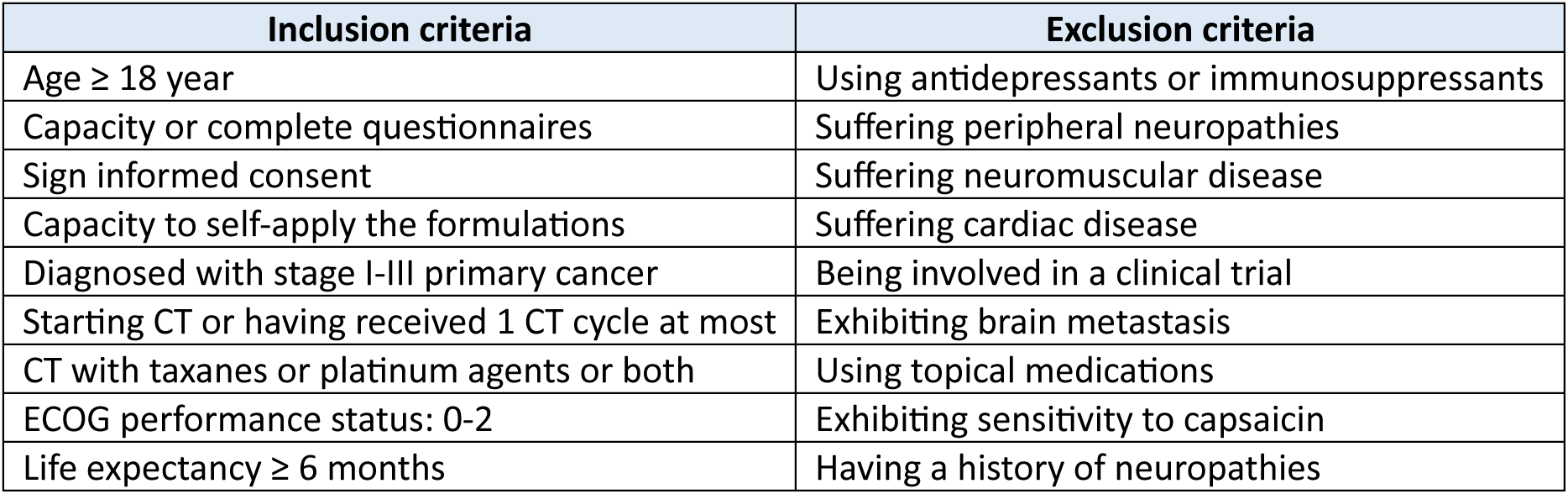
Inclusion-exclusion criteria used to recruit the patient cohort.

Once the informed consent was signed, a code was assigned to the patient and provided with a moisturizing formulation (labelled as PB-011) or the nociceutical formulation (labelled as PB-012) according to the randomization protocol.

### Randomization and blinding

Patients were randomized using a computer-generated random number sequence with a block size of ten. Randomization was stratified according to the study center. Treatment allocation was blind to study investigators and patients.

### Procedures and assessments

The study protocol established that eligible patients must have a chemotherapeutic treatment containing taxanes and/or platinum agents, in combination with other agents as considered by study investigators. Recruited patients were instructed to apply the assigned ointment onto the hands once daily at the beginning of the prescribed chemotherapeutic treatment, and to inform of any sensory discomfort (tingling, itching, altered sensation to cold or heat) that appear in hands and/or feet throughout CT treatment. Because 20-30% of patients treated with taxanes or platinum agents do not develop the neuropathy, the study protocol was designed to identify the group of patients that do not get CIPN in both arms.

The primary endpoint was the percentage of patients that were free of CIPN during CT. The secondary endpoint was the percentage of CT cycles free of neuropathic symptoms. We also evaluated the percentage of patients that experienced a reduction in the severity of the neuropathic symptoms. The compliance with QT treatment was also recorded, including dose reduction, CT cessation, and tolerance to the creams.

The onset of CIPN was diagnosed by study investigators using the most recent version of the National Cancer Institute’s Current Toxicity Criteria (CTC AE) version 5.0. After the onset of sensory symptoms in the hands and/or feet, patients were indicated to start applying the assigned formulation onto their hands and feet twice daily for up to one month after the end of prescribed chemotherapy treatment. These patients completed the Leonard Scale Questionnaire ^22^ to every three weeks until one month after the end of treatment to assess the severity of the CIPN sensory symptoms according of how much the symptoms affected their daily activities. The severity of the sensory symptoms was divided as hardly at all bothered (0-1), moderately bothered (2), and extremely bothered (≥3) as indicated by Leonard et al.^22^.

### Topical formulations

Moisturizing and nociceutical formulations were provided by the sponsor in identical white tubes, labelled as PB-011 and PB-012. Both creams had the same appearance and similar consistency and odor. Moisturizing cream incorporated Capric Triglyceride, Glyceryl Behenate, Glyceryl Stearate, Hydrogenated Castor Oil, Diethylene Glycol Monoethyl Ether, and Tocopherol; the nociceutical formulation additionally incorporated Hydroxymethoxyiodobenzyl Glycolamide Perlargonate (INCI name of Calmapsin^®^).

### Adverse effects

Adverse events (AEs) associated to the formulations were assessed by the study investigators at follow-up visits and recorded in the data collection booklet. AEs were reported regardless of whether they were considered treatment-related by the investigator. AEs were classified according to CTCAE v.05. Dose reductions were permitted to manage adverse effects provoked by the CT.

### Statistical analyses

The estimated sample size was 120 participants (60 per arm). Sample size was estimated using the Wilcoxson-Mann-Whitney test (means, two groups), two-tails, with an effect size d of 0.60 for delaying and attenuating CIPN during CT cycles. The α level was set to 0.05 and the Power to 0.95 using the G*Power 3.1 software program. We considered a dropout of 20%, which inflated the sample size up to 140 patients (70 per arm).

Differences between means of the percentage of chemotherapy treatment CIPN free were analyzed with the non-parametric Mann-Whitney rank sum test. Differences between groups of categorical (yes/no or worse/better) or qualitative variables (none (0), mild (1-2) or severe (3-4)) of the Leonard Scale Questionnaire or body areas affected (hands/feet) were analyzed using the two-side Fisher’s exact test or Chi-square test. The α level was set to 0.05. Statistical analysis was performed with the Graphpad Prism 10.

## RESULTS

### Patients

A total of 142 cancer patients, 127 women (90%) and 15 men (10%) with mean age of 56±14 years, met the inclusion and exclusion criteria and were included in the study (Fig. 1, Tables 1 and 2). These patients were recruited between September 2022 and May 2024 and randomized into two groups, 70 were assigned to the moisturizing formulation and 72 to the nociceutical formulation. Among these, 88% and 76% (moisturizing vs. nociceutical) were diagnosed with breast cancer, 13% and 15% with gastrointestinal tumors, and 1.5% and 7% with gynecological cancer. A total of 72% and 74% of patients in both groups received taxanes (mainly paclitaxel)-based CT, being primarily patients with breast cancer, whereas 15% and 9% received platinum agents (mainly oxaliplatin) and 13 and 17% a combination of taxanes and platinum agents (Table 2).

**Figure 1.**
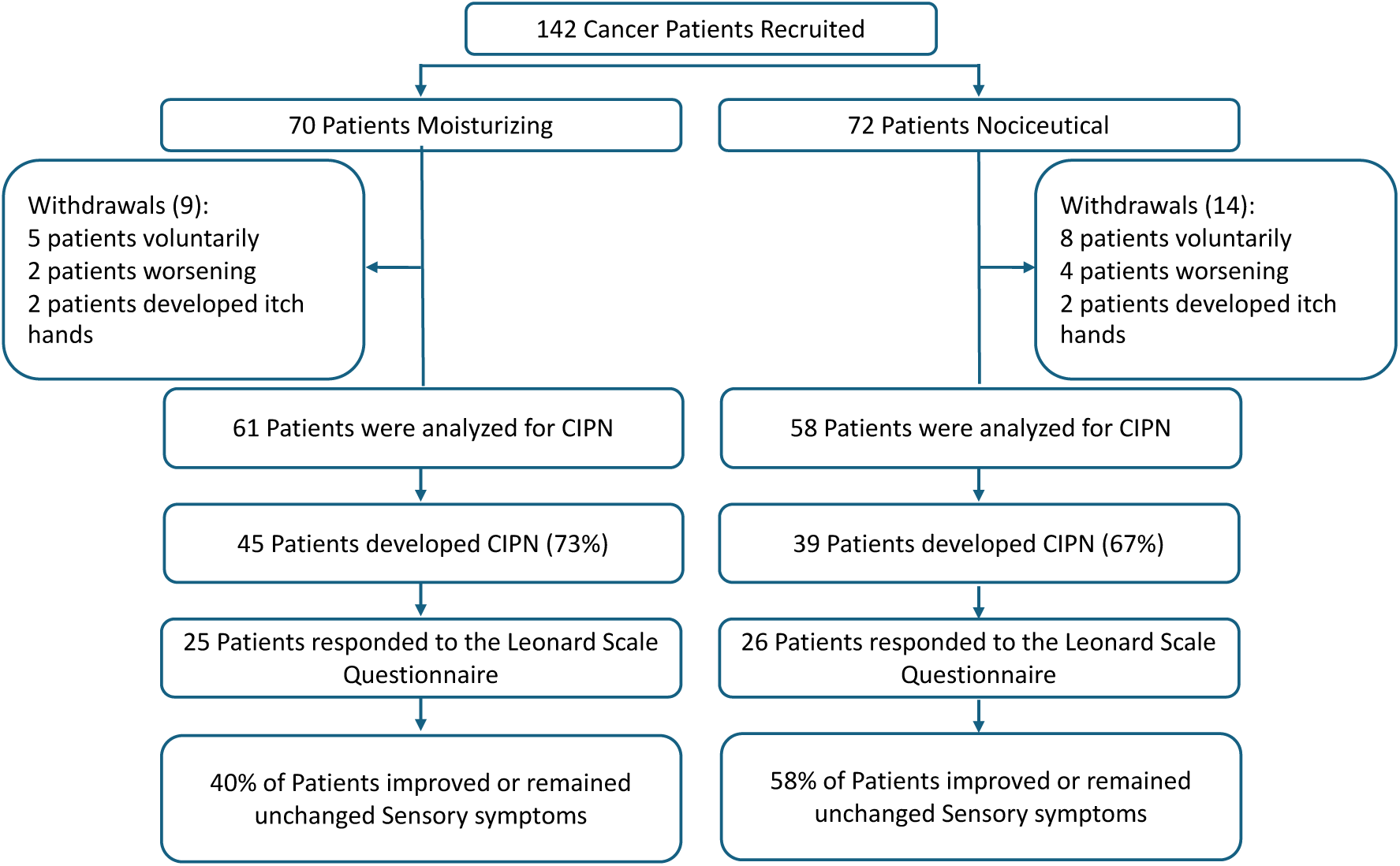
Consort diagram

**Table 2.**
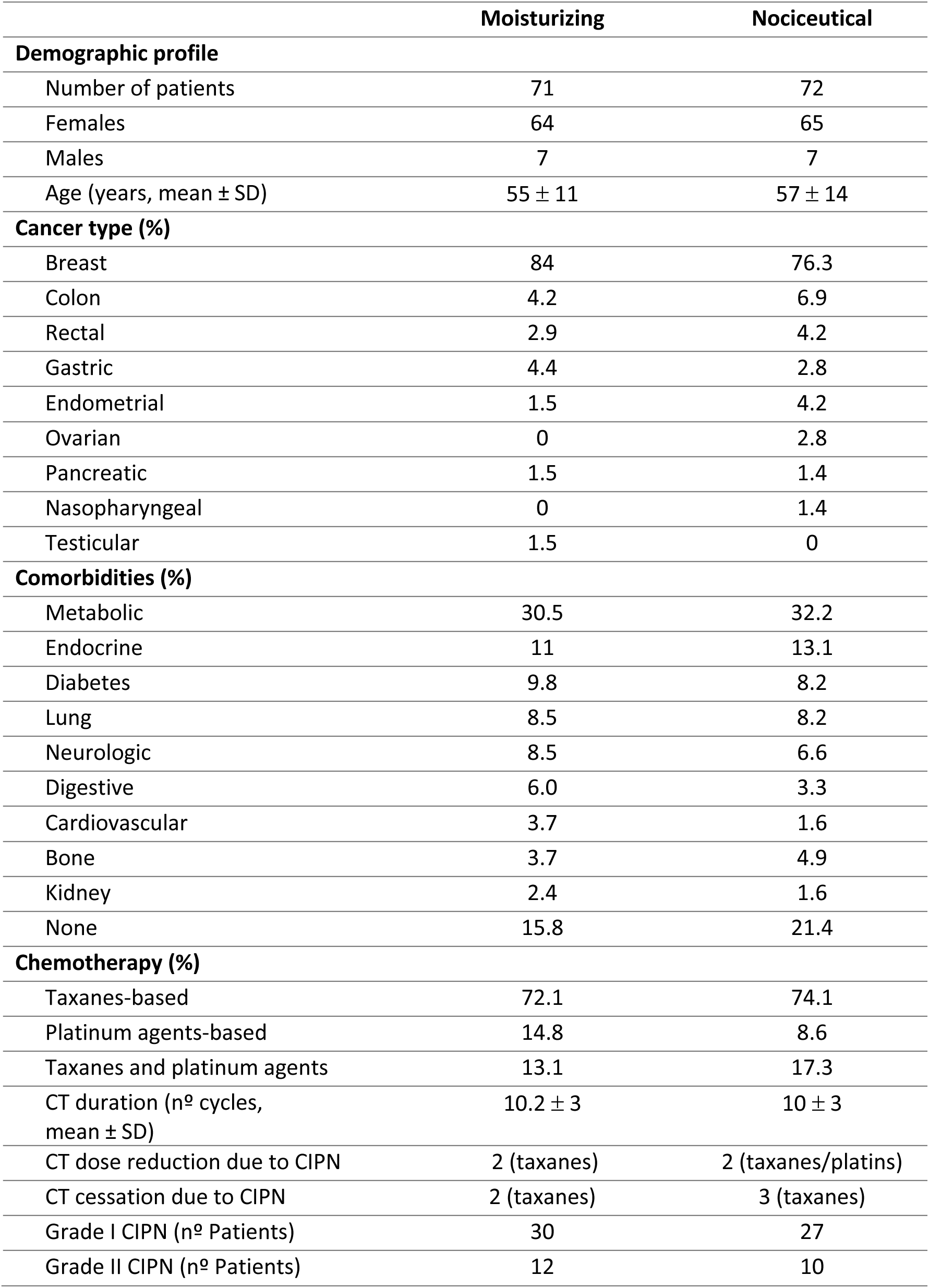
Cohort of patients.

|  | Moisturizing | Nociceutical |
| --- | --- | --- |
| **Demographic profile** |  |  |
| Number of patients | 71 | 72 |
| Females | 64 | 65 |
| Males | 7 | 7 |
| Age (years, mean ± SD) | 55 ± 11 | 57 ± 14 |
| **Cancer type (%)** |  |  |
| Breast | 84 | 76.3 |
| Colon | 4.2 | 6.9 |
| Rectal | 2.9 | 4.2 |
| Gastric | 4.4 | 2.8 |
| Endometrial | 1.5 | 4.2 |
| Ovarian | 0 | 2.8 |
| Pancreatic | 1.5 | 1.4 |
| Nasopharyngeal | 0 | 1.4 |
| Testicular | 1.5 | 0 |
| **Comorbidities (%)** |  |  |
| Metabolic | 30.5 | 32.2 |
| Endocrine | 11 | 13.1 |
| Diabetes | 9.8 | 8.2 |
| Lung | 8.5 | 8.2 |
| Neurologic | 8.5 | 6.6 |
| Digestive | 6.0 | 3.3 |
| Cardiovascular | 3.7 | 1.6 |
| Bone | 3.7 | 4.9 |
| Kidney | 2.4 | 1.6 |
| None | 15.8 | 21.4 |
| **Chemotherapy (%)** |  |  |
| Taxanes-based | 72.1 | 74.1 |
| Platinum agents-based | 14.8 | 8.6 |
| Taxanes and platinum agents | 13.1 | 17.3 |
| CT duration (nº cycles, mean ± SD) | 10.2 ± 3 | 10 ± 3 |
| CT dose reduction due to CIPN | 2 (taxanes) | 2 (taxanes/platins) |
| CT cessation due to CIPN | 2 (taxanes) | 3 (taxanes) |
| Grade I CIPN (nº Patients) | 30 | 27 |
| Grade II CIPN (nº Patients) | 12 | 10 |

The moisturizing and the nociceutical arms had 9 and 14 withdrawals respectively (Fig. 1), leading to 60 and 58 patients in each arm analyzed for the primary endpoint. Withdrawals were due to tumor progression and cessation of the prescribed CT (2 and 3 patients for moisturizing and nociceutical formulation, respectively), volunteer withdrawal (5 and 9 patients), and manifestation of itch upon application of the formulations (2 and 2 patients) (Fig. 1). The major comorbidities of these patients were metabolic (30% and 32%, mainly obesity and metabolic syndrome), endocrine (11% and 13%), and diabetes (10% and 8%), while 15% and 21% did not exhibited any (Table 2).

The Leonard Scale Questionnaire was answered by 25 patients and 26 patients of the moisturizing and nociceutical arms, respectively (Fig. 1).

### CIPN incidence during CT

Patients of both arms started using the formulations once a day in hands at the beginning of the prescribed CT. Application of the creams was monitored, on average, for 10±3 CT cycles as most patients were diagnosed with breast cancer and received taxanes. Both formulations were safe only exhibiting as AEs the manifestation of itch (grade 1, CTC AE v5.0) in hands upon application in 4 patients (2 in each arm). All other AEs were related with the CT and recorded by study investigators using the CTC AE v5.0.

Analysis of the results shows that 45 of patients who completed the study in the moisturizing arm developed CIPN. In the nociceutical arm 39 patients developed the neuropathy (Fig. 2). Among these patients 70% developed G1 CIPN (30 vs 27), and 30% progressed to G2 CIPN (12 vs 10). CT dose reduction was needed in 2 patients in each arm due to CIPN, while CT cessation was required in 2 and 3 patients in the moisturizing and nociceutical arms, respectively (Table 2). Patients receiving taxanes or taxanes and platins exhibited a similar CIPN incidence in both groups (70% on average, Fig. 2B), while patients receiving platins (FOLFOX, CAPOX or XELOX) showed a 68% and 40% for moisturizing and nociceutical creams. Interestingly, we observed that taxanes promoted CIPN in hands and feet, while platinum agents primarily produced CIPN in hands. Nevertheless, this observation should be taken cautiously as the number of patients receiving only platins is low, not allowing for proper statistical analysis.

**Figure 2.**
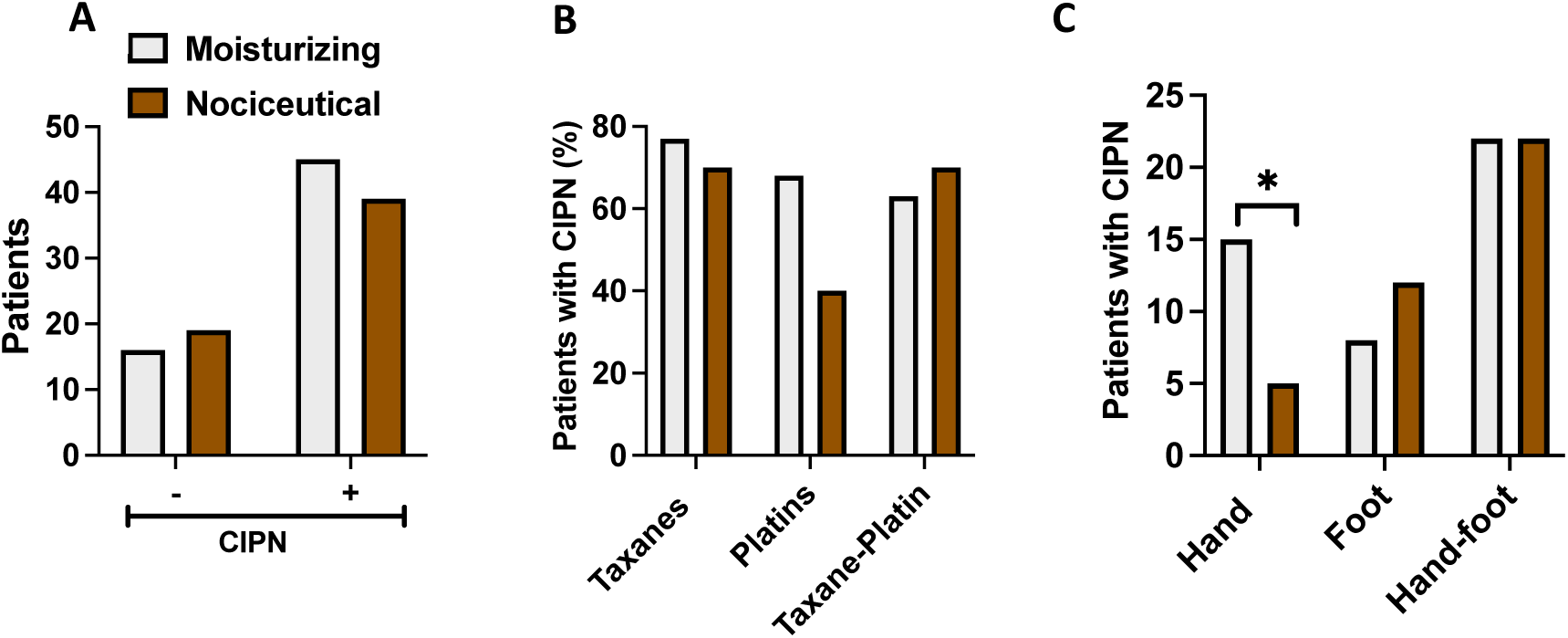
The use of the nociceutical formulation prevents CIPN in hands. **(A)** Percentage of patients that developed CIPN symptoms in both arms. (**B**) Percentage of patients that received taxanes and platinum agents that developed the neuropathy. (**C**) Percentage of patients that developed the neuropathy in hands, feet and hand&feet. Patients started to use the formulation once daily in hands at the beginning of the CT. Upon CIPN manifestation in feet or hands, patients applied the formulation twice daily. Data represent the percentage of patients that developed CIPN at any CT cycle of the 12 cycles considered in this study. For each arm, patients were normalized by the total cohort analyzed for CIPN, excluding patients that withdrawn the study. Statistical analysis was performed with the Fisheŕs exact test, *p=0.0307, 95% CI from 0.3438 to 0.9514, OR 0.2857. Only significant differences are indicated.

Although a tendency to an overall lower CIPN incidence was observed in the nociceutical arm, it did not reach statistical significance (73% vs 67%, p=0.10, two-tailed Fisheŕs exact test, Figs. 2A and 2B), possibly because of the small sample size. Analysis of patients that developed the neuropathy in hands (including hands only and hands & feet) also revealed a lower tendency in the nociceutical arm that did not reach statistical significance (57.1% vs. 42.9%, p=0.0783, 95% Confidence interval (95%CI) from 0.4293 to 1.0790, Odds Ratio (OR)=0.5, two-tailed Fisheŕs exact test). However, segregation of these patients into two groups (hands only and hands & feet) reveal a significant decrease in CIPN incidence in the group of hands only who used the nociceutical formulation from the beginning of CT (32% vs. 13%, p=0.0307, 95%CI from 0.3438 to 0.9514, OR=0.2857, two-tailed Fisheŕs exact test) (Fig. 2C). Taken together, these results suggest that the use of the nociceutical formulation from the beginning of the CT treatment reduced mainly the incidence of CIPN in hands of patients that developed only palmar CIPN.

### CIPN onset during CT cycles

We next evaluated if the use of the nociceutical formulation delayed the onset of CIPN sensory symptoms. For this analysis, we followed the percentage of patients who did not developed CIPN in each CT cycle throughout the treatment (Fig. 3A). As depicted, patients using the nociceutical formulation developed the neuropathy at later CT cycles than those using the moisturizing formulation, suggesting that the nociceutical cream delayed the appearance of the neuropathy. This analysis also revealed a tendency to a lower number of patients that did not develop CIPN during the 12 CT cycles in the nociceutical arm (42% and 34% for nociceutical vs moisturizing at cycle 12, respectively).

**Figure 3.**
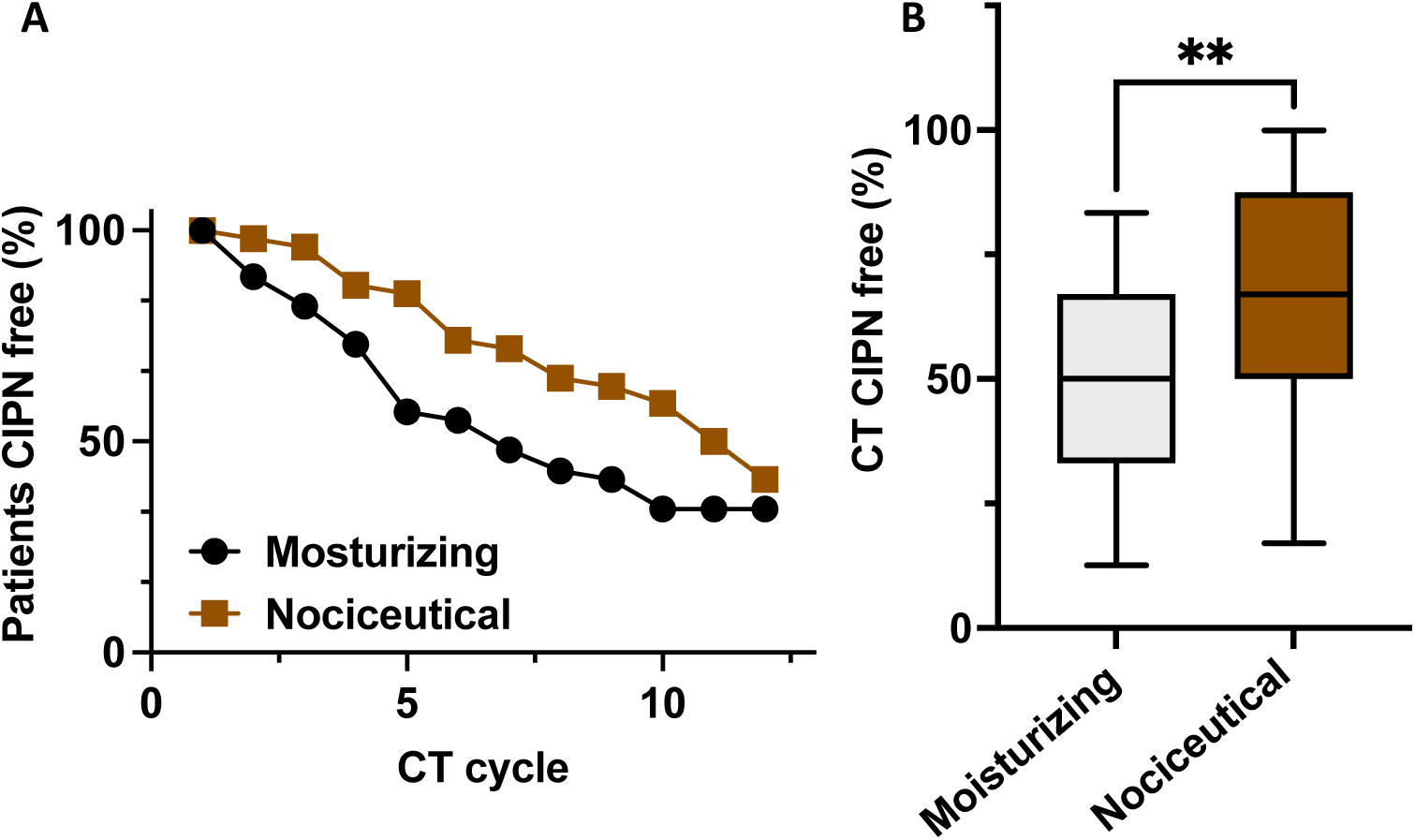
The nociceutical formulation delays the onset of CIPN as compared with the moisturizing formulation. **(A)** Percentage of patients CIPN free throughout the 12 chemotherapy cycles. (**B**) Percentage of the CT that patients developing CIPN are free of sensory symptoms. Data are mean ± SD, with n=42 and 37 patients for the moisturizing and nociceutical arms respectively (Fig. 1). Statistical analysis was performed using the non-parametric Mann-Whitney Rank sum test. **p=0.0091 (Sum or Ranks 651 and 1002; U=245; 95% CI from 33 to 63 and 50 to 83).

To statistically analyze the onset of CIPN appearance in patients developing the neuropathy, we estimated the mean time to display the neuropathy as the percentage of CT cycles that patients were free of neuropathic symptoms. Figure 3B shows that the nociceutical formulation deferred the onset of CIPN up to 35% of the CT cycles. While patients in the moisturizing arm exhibited 49±22% (mean ± SD) of CT cycles free of CIPN, patients of the nociceutical arm remained 65±27% (mean ± SD) of the CT cycles without neuropathic symptoms (p=0.0096, two-tailed Mann-Whitney Rank Sum Test) (Fig. 3B). Taken together, these results suggest that the use of the nociceutical formulation delayed the onset of CIPN during CT, being patients longer free of neuropathic symptoms.

### Impact of CIPN sensory symptoms

We utilized the Leonard Scale Questionnaire to investigate if the use of the nociceutical formulation, as compared to the moisturizing formulation, reduced the severity of CIPN sensory symptoms ^22^, improving the QoL of patients. The questionnaire was answered by 25 patients of the moisturizing group and 26 of the nociceutical arm (Fig.1). This survey revealed that 60% of patients using the moisturizing formulation frequently experienced CIPN bothering sensory symptoms as compared with 39% of patients using the nociceutical ointment (p=0.0017, 95%CI from 0.4709 to 0.8321, OR=0.39, two-tailed Fisheŕs exact test, Fig. 4A).

**Figure 4.**
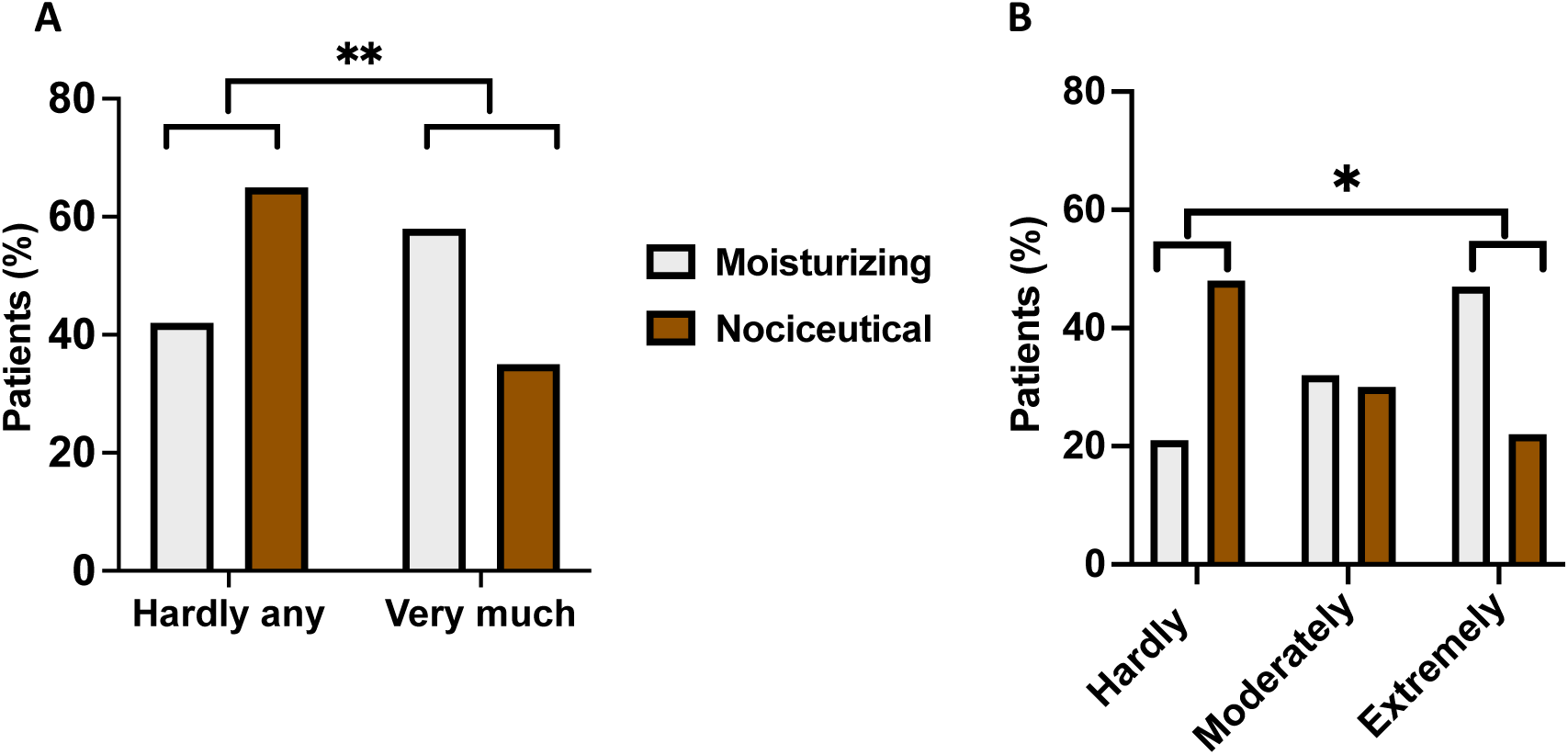
The use of the nociceutical formulation reduces the severity of the CIPN sensory symptoms. (**A**) Percentage of patients that indicate they experienced hardly any symptom or very much disturbing sensory symptoms in hands and/or feet. Statistical comparison was performed using the Fisheŕs exact test: **p=0.0017, 95% CI from 0.4709 to 0.8321, OR=0.39. N=23 and 25 for moisturizing and nociceutical formulations respectively (**B**) Percentage of patients indicating whether the sensory symptoms hardly, moderately or extremely bothered them performing daily activities. The severity of the sensory symptoms was divided as hardly at all bothered (0-1), moderately bothered (2), and extremely bothered (≥3) as indicated by Leonard et al.^22^. Statistical comparison was performed using the Contingency test Chi-square for trend, *p=0.0418. Only significant differences are indicated.

We further segregated patients according to how much the symptoms affected their daily activities. The severity the sensory symptoms was divided as hardly at all bothered, moderately bothered, and extremely bothered, according to the Leonard scale ^22^ (Fig. 4B). Analysis of these segregated data reveals a significative reduction in the extremely bothered group concomitant of an increase in the hardly at all bothered group (p=0.0418, Chi-square contingency test for trend). Thus, these data suggest that the topical use of the nociceutical formulation in hands and feet may reduce the severity of the neuropathic sensory symptoms and increase the QoL of patients that developed CIPN.

## DISCUSSION

CIPN is a disturbing side effect of most chemotherapy regimens that reduces the QoL of cancer patients by affecting their daily activities. This neuropathy may require a reduction, delay or even cessation of the CT, thus impacting patient therapeutic outcome. Increasing evidence supports that the neuropathy is primarily caused by a direct and accumulative excitation of distal peripheral nociceptive terminals by chemotherapeutic drugs that leads to an increment of the spontaneous activity of normally silent nociceptive endings ^17^. This condition may be exacerbated by the concomitant action of cutaneous, immune and Schwann cells that are also targeted by chemotherapeutics ^16^. Preclinical findings from different laboratories are signaling to thermosensory ion channels along with sodium channels as drivers of CIPN-mediated nociceptive excitability ^23,24^, suggesting that modulating their activity may protect epidermal terminals from the deleterious effects of chemotherapeutic drugs^5^. Here, we have tested this hypothesis and evaluated the effect of topically applying a nociceutical formulation containing a non-pharmacological ingredient that softly interacts with thermosensory nociceptive channels modulating the incidence, onset and severity of the neuropathy in a proof-of concept clinical study. Noteworthy, we observed a lower incidence on palmar CIPN in patients that applied the nociceutical formulation in hands since the starting of the CT treatment as compared with those using the moisturizing base formulation. Furthermore, this pilot study suggests a tendency reducing the overall CIPN incidence in the arm using the nociceutical formulation from the beginning of the CT treatment, although these data did not reach statistical significance most likely because of the limited cohort or/and dosage used. Indeed, it is possible that an increase in the number of topical applications (2-3) from the beginning may be also useful to protect epidermal nociceptive ends contributing to reduce hand-foot CIPN incidence. Thus, a confirmatory clinical study is needed to address these issues and determine if this strategy reduces the incidence of the neuropathy.

The secondary end point was reached as well. Our results show that the use of the nociceutical formulation significantly delayed the appearance of neuropathic symptoms to later CT cycles. Likewise, patients of the nociceutical arm tended to express milder neuropathic sensory symptoms, which bothered less their daily activities, thus experiencing a better QoL. Therefore, this proof-of-concept study reveals that the daily use of the nociceutical formulation delayed the appearance of the neuropathy and increase the QoL of patients.

The moisturizing and nociceutical formulations were safe, virtually free of AEs, only reporting pruritus grade 1 in 2 patients in each arm upon application of the formulations in hands. Both the moisturizing and nociceutical formulations were anhydrous ointments due to the water-sensitive ester-bond incorporated into the soft nociceutical ingredient Calmapsin^®^. An anhydrous basic formulation has the advantage of not requiring any preservative nor antimicrobial components that may not be tolerated by cancer patients treated with CT. In this regard, the itchy reaction observed may be related to the presence of ethoxydiglycol (Transcutol^®^) in the formulations ^25^. This alcohol is used in topical formulations to increase the permeability through the stratum corneum but may produce a sensitivity response in some individuals either directly or by increasing the skin permeability to other potential antigens. As Calmapsin^®^ is a hydrophobic molecule that readily permeates through the stratum corneum ^20^, the presence of ethoxydiglycol in the formulation may be adjusted in the nociceutical formulation.

CIPN is currently treated with antidepressants or anticonvulsants drugs, such as duloxetine or gabapentin, and Qutenza^®^ patches that provide relieve to patients although both treatments exhibit side-effects that limit their use and are primarily applied to grades III/IV of the neuropathy ^5^. Drugs such as lidocaine, cannabinoids, ketamine and menthol are also considered ^5^, although the ASCO guidelines for the prevention and management of CIPN do not recommend the use of these products due to the lack of sufficient clinical evidence ^13^. Currently, several clinical studies are in progress evaluating the efficacy and safety of different strategies including the administration of metformin and IL-6, as well as exercise, nutrition and acupuncture as registered in the NIH clinical trials web (https://clinicaltrials.gov/). Notably, sensorimotor exercise and resistance training appears to have a beneficial effect preventing the appearance of CIPN^26^. Additionally, clinical studies testing the neuroprotective glycosphingolipid monosialotetrahexoxylganglioside (GM1), administered intravenously (40 to 80 mg/daily), suggest a protective effect on acute and chronic CIPN^27,28^, although further studies are required as a Phase III clinical trial did not observe a preventive effect on oxaliplatin-induced peripheral neuropathy ^9^. Interestingly, GM1 has been shown that modulates the activity of TRPV1 channels^29,30^, implying that part of its protective activity may mediated through modulation of thermosensory nociceptor channels akin to our nociceutical formulation.

Complementing these strategies, we reasoned that soft compounds acting on the epidermis and hydrolyzed in the dermis into metabolites that are readily eliminated, would be an appropriate strategy to enhance the efficacy protecting epidermal sensory endings, while preventing the systemic distribution of the nociceutical and any potential interfere with CT. In addition, as dysesthesia and paresthesia are primarily caused by excitation of epidermal nociceptor endings, a topical action would have a more selective and safe action as compared with a systemic administration. We selected as soft TRPV1 modulator (Calmapsin^®^) that exhibits mild cross reactivity with other thermosensory drivers of nociceptor excitability, i.e. TRPA1 and TRPM8^21^. In addition, Calmapsin^®^ may synergistically act with the moisturizing components used in the formulation, as lipids are good modulator of nociceptive channels, receptors and membrane enzymes^31–33^, and tocopherol exhibits antioxidant actions. Thus, the biological action exerted by our nociceutical formulation, with multitarget synergistic actions appears to be advantageous to protect epidermal nociceptive endings from the action of two chemotherapeutic drugs that exhibit different cytostatic mechanisms such as taxanes and platinum agents ^24,34,35^.

It should be noted that this pilot clinical study has some limitations that should be considered. First, our cohort of patients is skewed towards women with breast cancer treated with taxanes, having an underrepresentation of CIPN caused by platinum agents. Second, the limited sample size of both arms does not allow to properly segregate patients according to sex or comorbidities such as diabetes that also promote a peripheral neuropathy. Third, the cohort that answered the Leonard scale questionnaire is small, and the conclusions should be considered as preliminary. In this regard, this questionnaire should be complemented with the European Organization for Research and Treatment of Cancer Quality of Life Questionnaire EORTC-CIPN20 that evaluates physical, emotional and cognitive functions that help to identify better strategies to prevent and treat CIPN^5,36,37^. Fourth, alterations in the regularity of CT cycles due to secondary effects of the treatment may have impacted the onset of the CIPN. Despite these limitations, our findings suggest that protecting distal epidermal nociceptor endings could be a useful strategy for supportive care and quality of life of cancer patients that merits further investigation. Furthermore, it remains to be investigated if the use of a nociceutical formulation also prevents a late development of the neuropathy or its recurrence upon CT termination.

## CONCLUSION

In conclusion, our proof-of-concept clinical study suggest that topical use of a non-pharmacological nociceutical formulation containing a soft neuroprotective compound (named Oncapsisens®), along with fatty acids and tocopherol, during CT delays the onset of CIPN and lessens its severity, increasing the QoL as patients are less bothered by the neuropathic symptoms. These findings support the potential benefit of a topical strategy directed to protect epidermal nociceptive terminals to ameliorate the neuropathic symptoms of CIPN. They warrant the design of a larger, confirmatory, controlled clinical study to corroborate the clinical relevance and utility of our nociceutical approach to prevent or delay CIPN and/or lessen its impact on QoL.

## Supporting information

Study protocol

## Data Availability

Data will be shared upon request to Antonio Ferrer-Montiel.

## DATA SHARING STATEMENT

Data will be shared upon request to Antonio Ferrer-Montiel.

## AUTHORS CONTRIBUTIONS

Conception and design: AFM, CFH, MGE, AFC

Financial support: All authors

Administrative support: All authors

Collection and assembly of data: All authors

Data analysis and interpretation: All authors

Manuscript writing: All authors

Final approval of manuscript: All authors

Accountable for all aspects of the work: All authors

## ACKNOWLEDGEMENT

The authors would like to thank Dr. Javier Gallego, Alfredo Carrato and Yolanda Escobar for reviewing the manuscript and providing insightful discussions on the data.

