## Supplementary material for "A Non-Pharmacological, Nociceutical Formulation Lessens Chemotherapy-Induced Peripheral Neuropathy in Cancer Patients": Study protocol

### Protocol for All Interventional Studies Involving Human Subjects

#### Selection of patients, including both eligibility and ineligibility criteria

This was a multicenter, double-blind, randomized, placebo-controlled research clinical study was performed in 9 hospitals. The Ethics Committees of all participating Hospital approved the protocol and the informed consent documents. At each hospital, medical oncologists acting as principal investigators were responsible for:

(i) recruiting patients.
(ii) checking that they met the inclusion and exclusion criteria (Table below).
(iii) obtaining the signed informed consent.
(iv) completing the data collection booklet.
(v) and collecting the appearance of CIPN and CIPN grade, along with patient follow up.

Once the informed consent was signed, a code was assigned to the patient and provided with a moisturizing formulation (labelled as ointment A) or the nociceutical formulation (labelled as ointment B) according to the randomization protocol. Patients were randomized using a computer-generated random number sequence with a block size of ten. Randomization was stratified according to the study center. Treatment allocation was blind to study investigators and patients.

| Eligibility criteria | Ineligibility criteria |
| --- | --- |
| Age ≥ 18 year | Using antidepressants or immunosuppressants |
| Capacity or complete questionnaires | Suffering peripheral neuropathies |
| Sign informed consent | Suffering neuromuscular disease |
| Capacity to self-apply the formulations | Suffering cardiac disease |
| Diagnosed with stage I-III primary cancer | Being involved in a clinical trial |
| Starting CT or having received 1 CT cycle at most | Exhibiting brain metastasis |
| CT with taxanes or platinum agents or both | Using topical medications |
| ECOG performance status: 0-2 | Exhibiting sensitivity to capsaicin |
| Life expectancy ≥ 6 months | Having a history of neuropathies |

#### Schema and treatment plan, including administration schedule

The study protocol established that eligible patients must have a chemotherapeutic treatment containing taxanes and/or platinum agents, in combination with other agents as considered by study investigators. Recruited patients were instructed to apply the assigned ointment onto the hands once daily at the beginning of the prescribed chemotherapeutic treatment, and to inform of any sensory discomfort (tingling, itching, altered sensation to cold or heat) that appear in hands and/or feet throughout CT treatment.

The primary endpoint was the percentage of patients that were free of CIPN during CT. The secondary endpoint was the percentage of CT cycles free of neuropathic symptoms. We also evaluated the percentage of patients that experienced a reduction in the severity of the neuropathic symptoms. The compliance with QT treatment was also recorded, including dose reduction, CT cessation, and tolerance to the creams.

**Rules for dose modification.**

As indicated, recruited patients were instructed to apply the assigned ointment onto the hands once daily at the beginning of the prescribed chemotherapeutic treatment. After the onset of sensory symptoms in the hands and/or feet, the dose was augmented to twice daily in hands and feet for up to one month after the end of prescribed chemotherapy treatment. The onset of CIPN was diagnosed by study investigators using the most recent version of the National Cancer Institute's Current Toxicity Criteria (CTC AE) version 5.0.

No other dose modifications were allowed during the study.

Modification of CT was determined by study investigators according to National Cancer Institute's Current Toxicity Criteria (CTC AE) version 5.0.

**Measurement of treatment effect including response criteria, definitions of response and survival, and methods of measurement**

The onset of CIPN was diagnosed by study investigators using the most recent version of the National Cancer Institute's Current Toxicity Criteria (CTC AE) version 5.0. After the onset of sensory symptoms in the hands and/or feet, patients were indicated to start applying the assigned formulation onto their hands and feet twice daily for up to one month after the end of prescribed chemotherapy treatment. These patients completed the Leonard Scale Questionnaire to every three weeks until one month after the end of treatment to assess the severity of the CIPN sensory symptoms according of how much the symptoms affected their daily activities. The severity of the sensory symptoms was divided as hardly at all bothered (0-1), moderately bothered (2), and extremely bothered (≥3) as indicated by *Leonard GD, Wright MA, Quinn MG, et al: Survey of oxaliplatin-associated neurotoxicity using an interview-based questionnaire in patients with metastatic colorectal cancer. BMC Cancer 5:116, 2005*

**Reasons for early cessation of trial therapy**

Patients stop using the creams if a related adverse effect arise or if considered by the principal investigator according to National Cancer Institute's Current Toxicity Criteria (CTC AE) version 5.0.

**Objectives and entire statistical section (including end points)**

The estimated sample size was 120 participants (60 per arm). Sample size was estimated using the Wilcoxson-Mann-Whitney test (means, two groups), two-tails, with an effect size d of 0.60 for delaying and attenuating CIPN during CT cycles. The $\alpha$ level was set to 0.05 and the Power to 0.95 using the G*Power 3.1 software program. We considered a dropout of 20%, which inflated the sample size up to 140 patients (70 per arm).

Differences between means of the percentage of chemotherapy treatment CIPN free were analyzed with the non-parametric Mann-Whitney rank sum test. Differences between groups of categorical (yes/no or worse/better) or qualitative variables (none (0), mild (1-2) or severe (3-4)) of the Leonard Scale Questionnaire or body areas affected (hands/feet) were analyzed using the two-side Fisher's exact test or Chi-square test. The $\alpha$ level was set to 0.05. Statistical analysis was performed with the Graphpad Prism 10.
